# Exploring Access to Social Protection for People with Disabilities in Bangladesh

**DOI:** 10.1101/2023.11.06.23298193

**Authors:** Mizanur Rahman, Md Shohel Rana, Md Mostafizur Rahman, Md Nuruzzaman Khan

## Abstract

**Purpose:** This study aimed to assess the accessibility of social protection programs for individuals with disabilities in Bangladesh and identify factors at the individual, household, and community levels influencing this accessibility.

**Methods:** We analyzed data from 4,293 respondents in the 2021 National Survey on Persons with Disabilities. We categorized participation in social protection programs as follows: no assistance (0), support received within six months (1), and support received beyond six months (2). Explanatory variables were considered at individual, household, and community levels. A multilevel multinomial logistic regression model assessed associations, with two age groups: 0-17 and 18+.

**Results:** Only 38% reported inclusion in social protection programs within six months, rising to 48% for support beyond six months. Disability allowances were the most common, followed by old age allowances and VGD/VGF assistance. Inclusion was likelier for older, unmarried, widowed, divorced, or separated individuals with disabilities. Conversely, those with higher education, wealthier households, and residing in Dhaka division were less likely to be included. Among children aged 0-17, multiple disabilities increased the likelihood of inclusion.

**Conclusion:** These findings underscore the urgent need for more comprehensive and inclusive social protection policies and programs to support the well-being of individuals with disabilities in Bangladesh.

## Introduction

Social protection programs, such as old age allowances and widow allowances, are commonly implemented in low-and middle-income countries (LMICs) by both government and non-government organizations (NGOs) [1]. These initiatives aim to provide a safety net for individuals and households facing specific needs due to poverty and other vulnerabilities, with the ultimate goal of enhancing their livelihoods [2, 3]. By doing so, the overarching objectives are to meet the basic needs of vulnerable populations, encompassing aspects such as health and education, while integrating them into mainstream society [2]. Predominantly, social protection programs in LMICs take the form of financial assistance or the provision of food, distributed to beneficiaries at fixed intervals, often on a monthly basis [4, 5]. Several research studies conducted across diverse settings have demonstrated the effectiveness of these social protection programs in improving the livelihoods of vulnerable populations, countering the common issues of mismanagement and corruption in beneficiary selection [6-8]. Consequently, these programs have been incorporated as a major target within the Sustainable Development Goals (SDGs, target 1.3) to be achieved by 2030, with the aim of reducing and preventing poverty and enhancing the livelihoods of vulnerable populations [9].

With 16% of the global population, individuals with disabilities constitute the world’s largest minority group [10]. Over 80% of them reside in LMICs, with their numbers steadily increasing due to factors such as improved survival rates among those born with disabilities and the rise in disability resulting from road traffic injuries and other causes [10-12]. Notably, a significant proportion of this population resides within the lower socio-economic segments, indicating that the majority of individuals with disabilities, especially those of younger ages, are living in poverty [13-15]. Moreover, their limited income-generating opportunities render them highly reliant on social protection programs [16, 17]. The prevalence of predominant socio-cultural norms in LMICs, which often attribute disabilities to curses and perceive them as permanent burdens, further exacerbates their situation [18, 19]. This situation is particularly pronounced in Bangladesh [7, 20], where approximately 3% of the total population is classified as disabled, and such misconceptions are widespread.

However, despite these challenges and the substantial number of individuals with disabilities in Bangladesh, it remains unclear what percentage of them are covered by existing social protection programs and what specific forms of support they receive [21, 22]. Furthermore, the types and varieties of social protection programs available for individuals with disabilities remain undocumented [20]. The limited estimates provided by the Ministry of Social Welfare of Bangladesh do not align with the true prevalence of disabilities and solely account for the support they offer [23]. NGOs also play a role in implementing social protection programs; however, their coverage remains undisclosed [24]. Additionally, the factors associated with inclusion in social protection programs are not well-understood in Bangladesh, as like other LMICs, as existing studies have reported conflicting associations due to small sample sizes and less precise data analysis methods [25-27]. This study aims to address these gaps by examining the coverage of social protection programs for individuals with disabilities and identifying the factors influencing their inclusion in these programs.

## Methods

### Sampling strategy

Data were obtained from the 2021 National Survey on Persons with Disabilities (NSPD) conducted by the Bangladesh Bureau of Statistics (BBS) [28]. The survey employed a two-stage stratified random sampling technique to identify the nationally representative households from where respondents were included. In the first stage, 800 primary sampling units (PSUs) were selected randomly from the list of 293,579 PSUs generated by the Bangladesh Bureau of Statistics. Household listing operation was then conducted in each selected PSU. Subsequently, 45 households were systematically chosen from each selected PSU in the second sampling stage. This approach yielded a roster of 36,000 households, of them data collected were undertaken in 35,493 households, attaining a coverage rate of 98.6%. There were 155,025 respondents in these selected households and all of them were included in the survey. Detailed description regarding the survey has been presented elsewhere [28].

### Analytical sample

We analyzed data from 4,293 respondents, which constitutes 2.79 percent of the total NSPD sample. The criteria for inclusion were: (i) persons with self-reported disability and (ii) respond to the questions related to social protection. The process of collecting disability related data are published elsewhere [28, 29].

### Outcome variables

The outcome variable considered was the respondents’ inclusion in any social protection program (yes, no). During the survey, participants were asked about their inclusion in any social protection programs run by the government and non-government organizations. They were provided a list to response along with an option to write if the supports they received are not already listed. It includes: disability allowances, education stipend for disabled, freedom freighter family allowances, old age allowances, widow allowances, maternity allowance, assistance-VGD/VGF, money/food for work and others allowance under the social safety net program. They were also asked to respond “no” if they are not part of any social protection programs. A separate question inquired about the timing of their receipt of support under any social protection programs. From these responses, we derived a variable with three categories: absence of social assistance (0), individuals who had received social support within six months of the survey (1), and those who had received social support at any point beyond six months (2).

### Explanatory variables

We used a comprehensive two-stage selection process to select explanatory variable. We first generate a list of potential explanatory variables based on the extensive review of the available relevant studies conducted in LMICs [12, 25, 30, 31]. Subsequently, these identified variables were cross-referenced with the survey data to verify their availability. Those variables that were available in the survey were subsequently classified according to the socio-ecological model of health into three tiers: individual level factors, households level factors, and community level factors. The individual-level factors encompassed the respondent’s age (children aged 0-17 years, adults aged 18- 59 years and older aged 60 and above), educational attainment (no education, primary, secondary, and higher), gender (male or female), occupation (agriculture, blue-collar work, pink-collar work, white-collar work, student, housewife, and others), marital status (married, unmarried, or widowed/divorced/separated) and religion (Islam, others). At the household level, a variable representing wealth quintiles of household was included (poorer, poorest, middle, richer, richest). This variable was formulated by the survey authority by using principal component analysis of household asset-related variables, such as roofing type and ownership of a refrigerator. Respondents’ place of residence (urban and rural) and region of residence (Barishal, Chattogram, Dhaka, Khulna, Mymensingh, Rajshahi, Rangpur, and Sylhet) were considered as community level variables.

### Statistical analysis

Descriptive statistics were used to explore the characteristics of the respondents. Chi-square test was used to examine the statistical significance of the variation in inclusion of social protection program across the considered explanatory variables. Multilevel multinomial mixed effect logistic regression model was used to access the determinants of inclusion of social protection programs. The reason for using multilevel multinomial modelling lies on the hierarchical structure of the NSPD data, where individuals were nested within a households and households were nested within a cluster. Previous research found multilevel modeling produce more precise results for hierarchical data than conventional simple logistic regression models [32]. We run two separate model by splitting total sample in two groups based on their ages: aged 0-17 and aged 18-95. The reason for this division lies in the varying coverage of social protection programs for children as opposed to adults. Multicollinearity was checked before running each model and if evidence of multicollinearity was found, the relevant variable was deleted and the model runed again. We also calculate likelihoods of inclusion of social protection program by disability types by using multilevel logistic regression models. Results are reported as adjusted Risk Ratio (aRR)/adjusted Odds Ratio (aOR) with their corresponding 95% Confidence Intervals (95% CI). All statistical analyses were carried out using STATA/SE 14.0 (Stata Corp LP, College Station, Texas, United States of America).

### Ethics approval

The survey we analyzed was reviewed and approved by the Ethics Committee of the Bangladesh Bureau of Statistics. We obtained de-identified data from the BBS through submitting a research proposal for this research. Therefore, there is no need for any additional ethical approval to conduct this study.

## Result

### Background characteristics of the persons with disability

The demographic characteristics of the respondents are presented in Table 1. The average age of the respondents was 41.4 years, with more than half of the total respondents being 18-59 years old. Nearly 59% of the total respondents were male. Around 56% of the total respondents were illiterate and nearly one third of total respondents identified them as unable to work. Over 80% of the total respondents resided in rural area while around 22% indicated Dhaka as their region of residence.

**Table 1:**
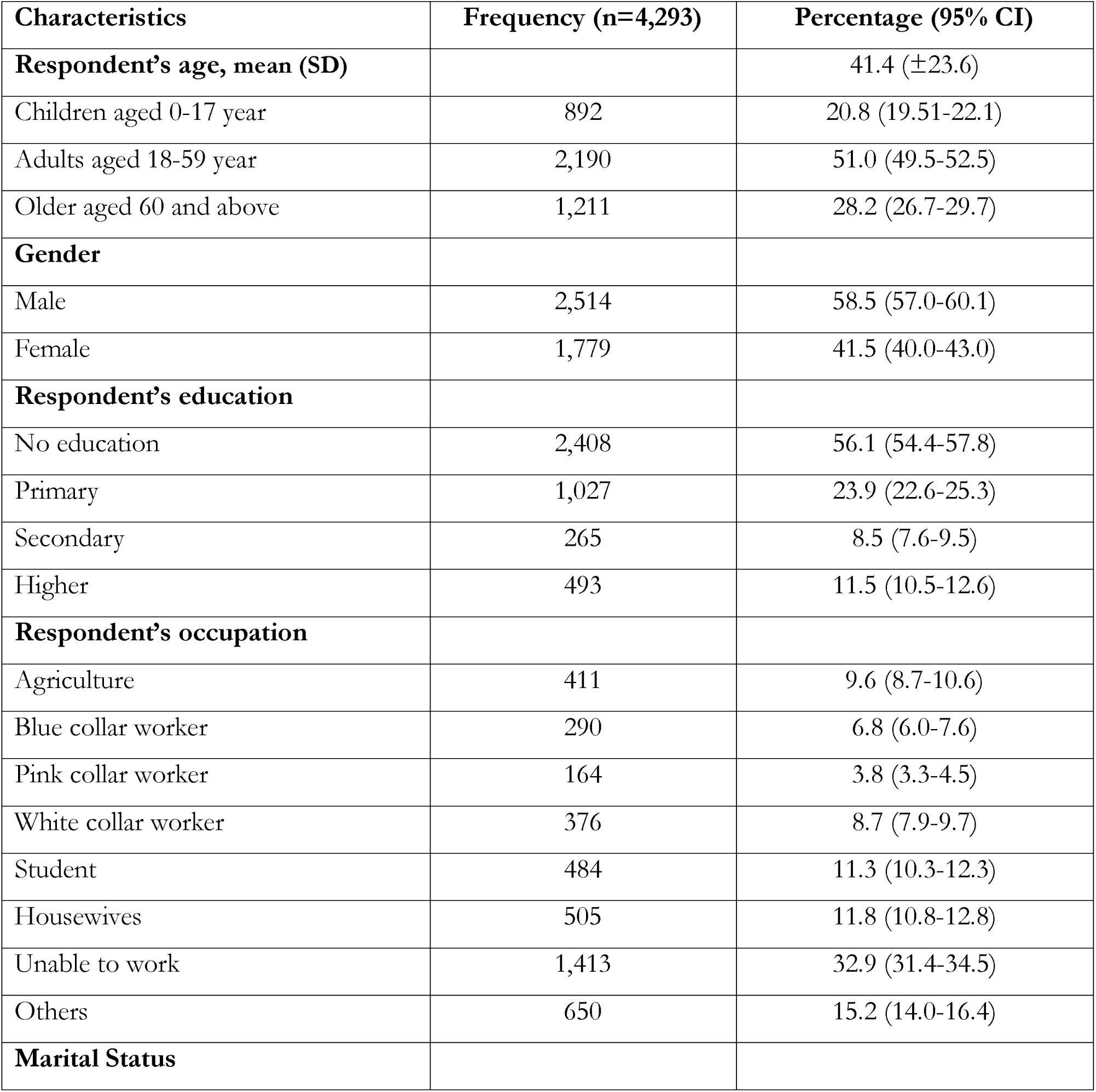
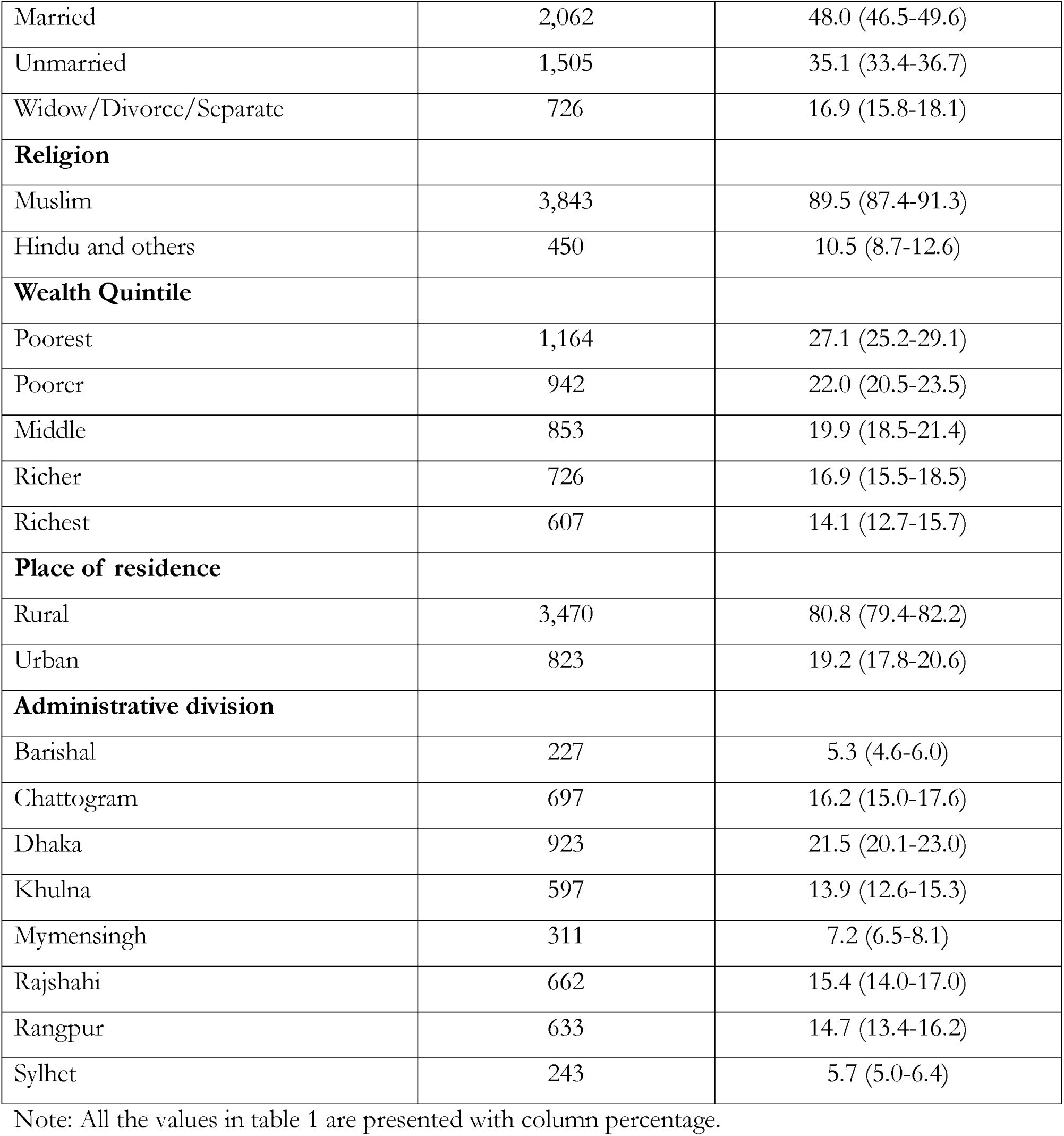
Background characteristics of the persons with disability in Bangladesh, 2021, N=4,293.

| **Characteristics** | **Frequency (n=4,293)** | **Percentage (95% CI)** |
| --- | --- | --- |
| **Respondent's age, mean (SD)** | | 41.4 (±23.6) |
| Children aged 0-17 year | 892 | 20.8 (19.51-22.1) |
| Adults aged 18-59 year | 2,190 | 51.0 (49.5-52.5) |
| Older aged 60 and above | 1,211 | 28.2 (26.7-29.7) |
| **Gender** |  |  |
| Male | 2,514 | 58.5 (57.0-60.1) |
| Female | 1,779 | 41.5 (40.0-43.0) |
| **Respondent's education** |  |  |
| No education | 2,408 | 56.1 (54.4-57.8) |
| Primary | 1,027 | 23.9 (22.6-25.3) |
| Secondary | 265 | 8.5 (7.6-9.5) |
| Higher | 493 | 11.5 (10.5-12.6) |
| **Respondent's occupation** |  |  |
| Agriculture | 411 | 9.6 (8.7-10.6) |
| Blue collar worker | 290 | 6.8 (6.0-7.6) |
| Pink collar worker | 164 | 3.8 (3.3-4.5) |
| White collar worker | 376 | 8.7 (7.9-9.7) |
| Student | 484 | 11.3 (10.3-12.3) |
| Housewives | 505 | 11.8 (10.8-12.8) |
| Unable to work | 1,413 | 32.9 (31.4-34.5) |
| Others | 650 | 15.2 (14.0-16.4) |
| **Marital Status** |  |  |
| Married | 2,062 | 48.0 (46.5-49.6) |
| Unmarried | 1,505 | 35.1 (33.4-36.7) |
| Widow/Divorce/Separate | 726 | 16.9 (15.8-18.1) |
| **Religion** |  |  |
| Muslim | 3,843 | 89.5 (87.4-91.3) |
| Hindu and others | 450 | 10.5 (8.7-12.6) |
| **Wealth Quintile** |  |  |
| Poorest | 1,164 | 27.1 (25.2-29.1) |
| Poorer | 942 | 22.0 (20.5-23.5) |
| Middle | 853 | 19.9 (18.5-21.4) |
| Richer | 726 | 16.9 (15.5-18.5) |
| Richest | 607 | 14.1 (12.7-15.7) |
| **Place of residence** |  |  |
| Rural | 3,470 | 80.8 (79.4-82.2) |
| Urban | 823 | 19.2 (17.8-20.6) |
| **Administrative division** |  |  |
| Barishal | 227 | 5.3 (4.6-6.0) |
| Chattogram | 697 | 16.2 (15.0-17.6) |
| Dhaka | 923 | 21.5 (20.1-23.0) |
| Khulna | 597 | 13.9 (12.6-15.3) |
| Mymensingh | 311 | 7.2 (6.5-8.1) |
| Rajshahi | 662 | 15.4 (14.0-17.0) |
| Rangpur | 633 | 14.7 (13.4-16.2) |
| Sylhet | 243 | 5.7 (5.0-6.4) |

Note: All the values in table 1 are presented with column percentage.

### Percentage distribution of inclusion in various types of social protection programs among individuals with disabilities in Bangladesh

Table 2 displays the different types of social protection programs through which individuals with disabilities have received social support. This includes those who received support within 6 months of the survey and those who received support at any time beyond six months. The average participation rate in any social protection program within the past 6 months was 37.7%, while with beyond six months, it was 47.4%. The majority of individuals who reported their inclusion in social protection programs mentioned receiving disability allowances, accounting for 69% of the cases. This was followed by recipients of old age allowances, constituting 16.3% of the total, and those who received assistance from the VGD/VGF program, amounting to 6.8%. A similar pattern emerged among respondents who reported receiving social support beyond six months. Roughly 60% of eligible disabled individuals, specifically those entitled to the freedom fighters’ allowance, reported not receiving such support. Similarly, approximately 57% of eligible disabled individuals did not receive the educational stipend.

**Table 2:** Social protection coverage among persons with disabilities in Bangladesh.

| Social protection program types | Total persons with disabilities |  |  |  | Distribution of social support uptake based on eligibility status |  |  |  |  |  |  |
| --- | --- | --- | --- | --- | --- | --- | --- | --- | --- | --- | --- |
|  | Individuals who had received social support within six months of the survey (n=1,620) |  | Individuals who had received social support at any point beyond six months (n=2,035) |  | Eligible respondents | No support (n=2,258) |  | Individuals who had received social support within six months of the survey (n=1,620) |  | Individuals who had received social support at any point beyond six months (n=415) |  |
|  | % | 95% CI | % | 95% CI |  | % | 95% CI | % | 95% CI | % | 95% CI |
| Disability allowances | 69.0 | 66.4-71.6 | 64.1 | 61.7-66.5 | 4,293 | 52.6 | 50.7-54.4 | 37.8 | 36.0-39.6 | 9.6 | 8.6-10.7 |
| Education stipend for disabled persons | 1.0 | 0.6-1.7 | 2.3 | 1.7-3.0 | 427 | 57.1 | 52.3-61.8 | 30.9 | 26.8-35.4 | 12.0 | 9.1-15.5 |
| Freedom fighter family allowances | 0.9 | 0.5-1.6 | 0.7 | 0.4-1.3 | 682 | 60.0 | 56.0-63.8 | 30.8 | 27.1-34.8 | 9.2 | 7.2-11.7 |
| Old age allowances | 16.3 | 14.4-18.3 | 15.3 | 13.7-17.0 | 1,362 | 48.3 | 45.2-51.3 | 42.1 | 39.2-45.1 | 9.6 | 8.1-11.5 |
| Widow allowances | 3.8 | 3.0-4.9 | 3.7 | 2.9-4.7 | 714 | 47.5 | 43.5-51.5 | 41.9 | 38.1-45.8 | 10.6 | 8.5-13.4 |
| Maternity allowance | - | - | 0.1 | 0.0-0.4 | 774 | 55.4 | 51.6-59.1 | 34.8 | 31.3-38.5 | 9.8 | 7.8-12.3 |
| Assistance-VGD/VGF | 6.8 | 5.5-8.5 | 9.0 | 7.7-10.5 | 4,292 | 52.6 | 50.8-54.5 | 37.7 | 35.9-39.5 | 9.7 | 8.7-10.7 |
| Money/Food for work | 1.1 | 0.7-1.8 | 2.7 | 2.0-3.5 | 2,916 | 50.8 | 48.6-53.0 | 39.1 | 37.0-41.3 | 10.1 | 8.9-11.5 |
| Others allowance | 1.1 | 0.6-1.7 | 2.2 | 1.6-3.1 | 3,933 | 51.5 | 49.7-53.5 | 38.5 | 36.6-40.3 | 10.0 | 8.9-11.1 |
| **Overall allowance acceptance** | **37.7** | **36.0-39.6** | **47.4** | **45.6-49.2** |  |  |  |  |  |  |  |

**Note:** We assessed eligibility based on the background characteristics of the respondents. For example, in the case of the freedom fighter family allowance, we examined the total number of eligible individuals with disabilities among families qualified to receive the freedom fighter allowance and how many of them reported receiving this allowance. \*Presented values are in row percentage.

The percentage distribution of none-inclusion of any social protection program by the children aged 0-17 are presented in Figure 1. Availability of no services at the residing area (41.0%) was found as most common reason followed by lack of information regarding the availability of services (17.2%), high service cost (16.4%) and negative attitude towards services (16.2%). We found male-female differences in responding reasons for not accessing social support. The data pertinent to adults aged 18 or older did not provide information on their reasons for not being included in any social protection program; therefore, we were unable to report on this matter.

**Figure 1:**
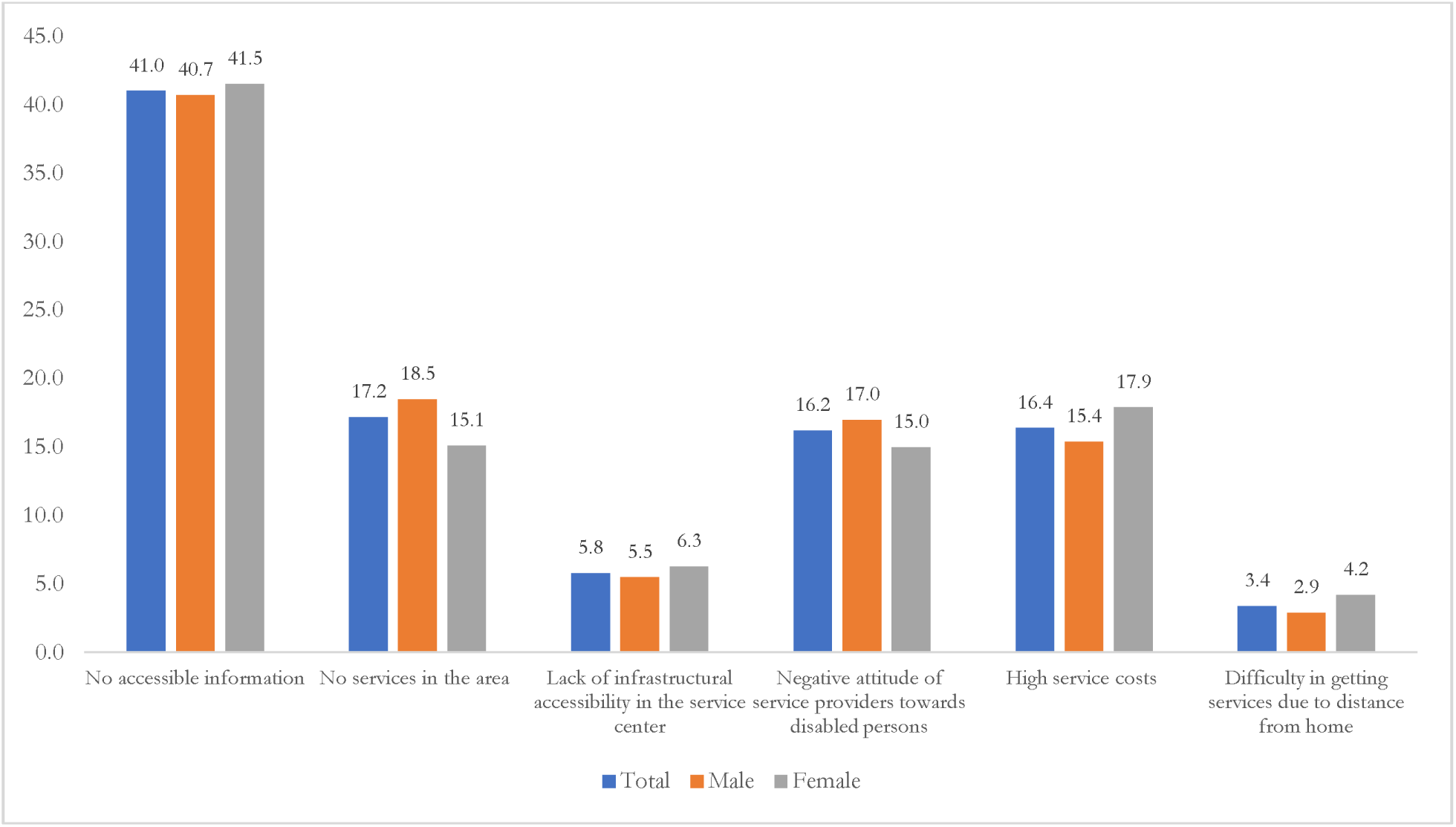
Reasons for not receiving benefits of social safety-net program among persons with disability aged 0-17 years.

### Distribution of inclusion of social protection program by persons with disabilities across individual, household and community level factors

Table 3 presents distribution of inclusion of social protection program by persons with disabilities across considered individual, households and community level factors. Persons with disabilities who aged 60 or older, had no formal education, was unable to work and those classified as widowed, divorced, or separated reported higher percentage of inclusion of social protection program. At the regional level, it was found that persons with disabilities who residing in the Mymensingh and Barishal divisions reported higher inclusion of social protection programs. We found statistically significant variations of inclusion of social protection program across considered individual, household, and community-level characteristics, except respondents’ gender and religion.

**Table 3:** Inclusion of social protection programs of persons with disability at anytime (n=4293)

| Characteristics | Inclusion of Social Protection Programs |  | P-value |
| --- | --- | --- | --- |
|  | Yes | No |  |
| **Individual level factor** |  |  | p<0.001 |
| **Respondent's age** |  |  |  |
| Children aged 0-17 year | 39.2 | 60.8 |  |
| Adults aged 18-59 year | 47.7 | 52.4 |  |
| Older aged 60 and above | 52.9 | 47.1 |  |
| **Gender** |  |  | p=0.278 |
| Male | 46.7 | 53.3 |  |
| Female | 48.4 | 51.6 |  |
| **Respondent's education** |  |  | p<0.001 |
| No education | 51.4 | 48.7 |  |
| Primary | 46.1 | 53.9 |  |
| Secondary | 38.8 | 61.2 |  |
| Higher | 37.1 | 63.0 |  |
| **Respondent's occupation** |  |  | p<0.001 |
| Agriculture | 47.5 | 52.6 |  |
| Blue collar worker | 42.2 | 57.8 |  |
| Pink collar worker | 44.6 | 55.5 |  |
| White collar worker | 51.4 | 48.6 |  |
| Student | 41.6 | 58.4 |  |
| Housewives | 40.7 | 59.3 |  |
| Unable to work | 55.0 | 45.0 |  |
| Others | 41.1 | 58.9 |  |
| **Marital Status** |  |  | p<0.001 |
| Married | 44.8 | 55.2 |  |
| Unmarried | 48.0 | 52.0 |  |
| Widow/Divorce/Separate | 53.5 | 46.5 |  |
| **Household level factor** |  |  |  |
| **Religion** |  |  | p=0.066 |
| Muslim | 46.8 | 53.2 |  |
| Hindu and others | 52.2 | 47.8 |  |
| **Wealth Quintile** |  |  | p<0.001 |
| Poorest | 52.5 | 47.6 |  |
| Poorer | 53.1 | 46.9 |  |
| Middle | 47.5 | 52.5 |  |
| Richer | 43.3 | 56.7 |  |
| Richest | 33.6 | 66.4 |  |
| **Community level factor** |  |  |  |
| **Place of residence** |  |  | p<0.001 |
| Rural | 49.4 | 50.6 |  |
| Urban | 39.0 | 61.0 |  |
| **Administrative division** |  |  | p<0.05 |
| Barishal | 51.9 | 48.1 |  |
| Chattogram | 46.7 | 53.3 |  |
| Dhaka | 41.5 | 58.5 |  |
| Khulna | 48.2 | 51.8 |  |
| Mymensingh | 54.5 | 45.5 |  |
| Rajshahi | 50.5 | 49.5 |  |
| Rangpur | 46.7 | 53.3 |  |
| Sylhet | 50.0 | 50.0 |  |

Note: All values in table 3 are presented with row percentage

### Individuals, households, and community level characteristics associated with inclusion of social protection program by the persons with disabilities

We conducted two separate multilevel multinomial models by dividing the total sample into two sub-groups: those aged 0-17 years and those aged 18 years or older. In the model for the 0-17 age group, we observed that a one-year increase in age was associated with a 1.30 times higher likelihood of inclusion in a social protection program for individuals who received social support within 6 months of the survey and for those who received social support at any time beyond six months. Furthermore, females with disabilities were more likely to report inclusion in social protection programs within 6 months of the survey (aRR, 2.0, 95% CI, 1.4-2.8) and at any time beyond six months, in comparison to persons with disabilities who were not part of any social protection program. We also identified a significant gradual decline in the likelihood of inclusion in social protection programs within 6 months of the survey as the level of education increased from no education. For instance, compared to illiterate persons with disabilities, inclusion in social protection programs decreased by 40% (aRR, 0.60, 95% CI, 0.4-0.9), 70% (aRR, 0.3, 95% CI, 0.2-0.5), and 80% (aRR, 0.2, 95% CI, 0.1-0.4) for disabled persons with primary, secondary, and higher education, respectively.

For persons with disabilities aged 18-95 years, a one-year increase in age was linked to 1 time increase in the likelihood of inclusion in social protection programs within 6 months of the survey and at any time beyond six months. However, the likelihood of inclusion in social protection programs within 6 months of the survey was found to be 20% lower among female persons with disabilities compared to male persons with disabilities (aRR, 0.8, 95% CI: 0.6-0.9). Additionally, we found that the likelihood of inclusion in social protection programs within 6 months declined by around 30-50% with the increase in the education level of persons with disabilities compared to those who were illiterate. There were higher probabilities of inclusion in social protection programs within 6 months among persons with disabilities working as white-collar workers (aRR, 1.6, 95% CI: 1.1-2.2), those unable to work (aRR, 1.5, 95% CI: 1.1-2.0), and students (aRR, 1.9, 95% CI: 1.1-3.3), in comparison to persons with disabilities engaged in agricultural work. As compared to married women, the likelihood of inclusion in social protection programs within 6 months was higher among unmarried (aRR, 2.5, 95% CI: 1.9-3.2) and widowed/divorced/separated (aRR, 1.4, 95% CI: 1.1-1.7) persons with disabilities. This relationship persisted for only unmarried persons with disabilities in terms of inclusion at any time beyond six months compared to their married counterparts. In contrast to persons with disabilities residing in poorer households, the likelihood of inclusion in social protection programs decreased with the increased of household wealth quintile in which persons with disabilities resided in. We observed a 40% lower likelihood of inclusion in social protection programs within 6 months of the survey among persons with disabilities residing in the Dhaka division as compared to those residing in the Barishal division.

**Table 4:** Factors associated with inclusion in social protection program by the persons with disabilities in Bangladesh.

| Characteristics | Children aged 0-17 |  | Adult and older aged 18-95 |  |
| --- | --- | --- | --- | --- |
|  | Individuals who had received social support within six months of the survey<br>aRR (95% CI) | Individuals who had received social support at any point beyond six months<br>aRR (95% CI) | Individuals who had received social support within six months of the survey<br>aRR (95% CI) | Individuals who had received social support at any point beyond six months<br>aRR (95% CI) |
| **Individual level factor** |  |  |  |  |
| **Respondent's age** | 1.3 (1.2-1.3) *** | 1.3 (1.2-1.4) *** | 1.0 (1.0-1.0) ** | 1.0 (1.0-1.0) |
| **Gender** |  |  |  |  |
| Male | 1.0 | 1.0 | 1.0 | 1.0 |
| Female | 2.0 (1.4-2.8) *** | 1.7 (1.0-2.9) ** | 0.8 (0.6-0.9) *** | 1.1 (0.8-1.5) |
| **Respondent's education** |  |  |  |  |
| No education | 1.0 | 1.0 | 1.0 | 1.0 |
| Primary | 0.6 (0.4-0.9) *** | 1.0 (0.5-1.7) | 0.7 (0.6-0.9) *** | 0.9 (0.7-1.2) |
| Secondary | 0.3 (0.2-0.5) *** | 0.3 (0.1-1.0) | 0.6 (0.4-0.7) *** | 0.4 (0.3-0.7) *** |
| Higher | 0.2 (0.1-0.4) *** | 0.6 (0.2-2.0) | 0.5 (0.4-0.6) *** | 0.6 (0.4-0.9) *** |
| **Respondent's occupation** |  |  |  |  |
| Agriculture | na | na | 1.0 | 1.0 |
| Blue collar worker | na | na | 0.9 (0.6-1.3) | 1.1 (0.7-1.9) |
| Pink collar worker | na | na | 1.2 (0.8-1.8) | 1.5 (0.8-2.7) |
| White collar worker | na | na | 1.6 (1.1-2.2) *** | 1.0 (0.5-1.7) |
| Student | na | na | 1.9 (1.1-3.3) ** | 2.3 (0.9-5.5) |
| Housewives | na | na | 1.2 (0.8-1.7) | 0.9 (0.5-1.5) |
| Unable to work | na | na | 1.5 (1.1-2.0) *** | 1.0 (0.6-1.5) |
| Others | na | na | 1.2 (0.9-1.7) | 1.6 (1.0-2.6) |
| **Marital Status** |  |  |  |  |
| Married | na | na | 1.0 | 1.0 |
| Unmarried | na | na | 2.5 (1.9-3.2) *** | 1.6 (1.0-2.4) ** |
| Widow/Divorce/Separate | na | na | 1.4 (1.1-1.7) *** | 1.0 (0.7-1.4) |
| **Household level factor** |  |  |  |  |
| **Religion** |  |  |  |  |
| Muslim | 1.0 | 1.0 | 1.0 | 1.0 |
| Hindu and others | 0.8 (0.4-1.5) | 0.9 (0.3-2.4) | 1.2 (1.0-1.5) | 1.4 (1.0-2.0) |
| **Wealth Quintile** |  |  |  |  |
| Poorest | 1.0 | 1.0 | 1.0 | 1.0 |
| Poorer | 1.3 (0.8-2.0) | 1.3 (0.6-2.7) | 1.0 (0.8-1.2) | 1.0 (0.7-1.4) |
| Middle | 0.9 (0.6-1.5) | 0.6 (0.2-1.5) | 0.8 (0.6-1.0) ** | 0.9 (0.7-1.3) |
| Richer | 1.3 (0.7-2.1) | 1.4 (0.6-3.3) | 0.7 (0.5-0.8) *** | 0.7 (0.5-1.0) ** |
| Richest | 1.0 (0.5-1.8) | 2.3 (1.0-5.6) | 0.4 (0.3-0.6) *** | 0.4 (0.2-0.6) *** |
| **Community level factor** |  |  |  |  |
| **Place of residence** |  |  |  |  |
| Rural | 1.0 | 1.0 | 1.0 | 1.0 |
| Urban | 0.9 (0.5-1.4) | 1.1 (0.5-2.4) | 0.8 (0.7-1.1) | 1.1 (0.8-1.6) |
| **Administrative division** |  |  |  |  |
| Barishal | 1.0 | 1.0 | 1.0 | 1.0 |
| Chittagong | 1.2 (0.5-2.7) | 1.6 (0.4-5.9) | 0.8 (0.6-1.2) | 0.7 (0.4-1.3) |
| Dhaka | 0.9 (0.4-2.1) | 1.2 (0.3-4.5) | 0.6 (0.4-0.9) *** | 0.9 (0.5-1.6) |
| Khulna | 1.5 (0.6-3.5) | 0.6 (0.1-3.0) | 0.9 (0.6-1.4) | 0.7 (0.4-1.3) |
| Mymensingh | 1.2 (0.5-2.9) | 0.9 (0.2-4.4) | 1.1 (0.7-1.7) | 1.0 (0.5-1.9) |
| Rajshahi | 1.5 (0.7-3.5) | 1.0 (0.2-4.0) | 0.9 (0.6-1.3) | 0.9 (0.5-1.6) |
| Rangpur | 1.3 (0.6-2.9) | 1.1 (0.3-4.7) | 0.8 (0.6-1.2) | 0.7 (0.4-1.3) |
| Sylhet | 1.1 (0.4-2.9) | 2.6 (0.6-10.9) | 0.8 (0.5-1.2) | 1.3 (0.7-2.6) |

Note: \*\*=p<0.05, \*\*\*=p<0.01, na= Not applicable

### Likelihood of inclusion of social protection program among various disability types

We conducted two additional adjusted multilevel analyses to assess the likelihood of enrollment in social support programs based on the type of disabilities, as presented in Table 5. Notably, we observed divergent patterns in the inclusion of social protection programs for individuals with disabilities aged 0-17 years and those aged 18 years or older. Among individuals aged 0-17 years, we found a notably higher likelihood of inclusion in social protection programs (adjusted odds ratio [aOR]: 2.6, 95% confidence interval [CI]: 1.2-5.9) for those who had multiple co-occurring disabilities compared to individuals with disabilities related to autism spectrum disorders. In contrast, for individuals aged 18 and older, we observed significantly lower odds of inclusion in social protection programs for those with mental illness (aOR: 0.1, 95% CI: 0.0-0.3), hearing impairment (aOR: 0.1, 95% CI: 0.0-0.6), other disabilities (aOR: 0.1, 95% CI: 0.0-0.7), and intellectual disabilities (aOR: 0.2, 95% CI: 0.0-0.6) when compared to individuals with disabilities related to autism or autism spectrum disorders.

**Table 5:** Adjusted likelihood of receiving social support allowance based on disability type.

| Type of disabilities | Disability among children<br>aged 0-17 years | Disability among adults and<br>older aged 18-95 |
| --- | --- | --- |
|  | aOR (95% CI) | aOR (95% CI) |
| Autism/Autism spectrum disorder | 1.0 | 1.0 |
| Physical disability | 0.9 (0.4-2.0) | 0.3 (0.1-1.1) |
| Mental illness disability | 0.4 (0.2-1.2) | 0.1 (0.0-0.3) *** |
| Visual impairment | 0.8 (0.3-2.0) | 0.3 (0.1-1.3) |
| Speech impairment | 1.1 (0.5-2.7) | 0.3 (0.1-1.2) |
| Intellectual disability | 0.6 (0.2-1.4) | 0.2 (0.0-0.6) *** |
| Hearing impairment | 0.6 (0.2-2.5) | 0.1 (0.0-0.6) *** |
| Hearing-Visual impairment | - | 0.2 (0.0-1.0) ** |
| Cerebral Palsy | 1.8 (0.8-4.4) | 0.8 (0.1-4.5) |
| Down syndrome | 0.4 (0.1-1.4) | 0.2 (0.0-1.0) ** |
| Multidimensional disability | 2.6 (1.2-5.9) *** | 0.4 (0.1-1.5) |
| Others | 0.2 (0.0-0.9) ** | 0.1 (0.0-0.7) *** |
| **Cluster-level variance (SE)** | 0.52 (0.30) | 0.67 (0.07) |
| **ICC** | 7.5% | 12% |
| **AIC** | 1044.497 | 4398.536 |
| **BIC** | 1188.635 | 4637.94 |

**Notes:** Models are adjusted for the individual, households and community level factors described in the explanatory variable section. \*\*= $p < 0.05$ , \*\*\*= $p < 0.01$

## Discussion

The study had two primary objectives: first, to assess the extent of inclusion of individuals with disabilities in social protection programs in Bangladesh, and second, to identify the factors associated with their inclusion in these programs. Only 38% of the persons with disabilities in Bangladesh reported being enrolled in any social protection program within six months of the survey, a percentage that rose to 48% when considering support received at any time beyond six months. Disability allowances were the most prevalent type of social protection program, followed by old age allowances and assistance within the VGD/VGF programs. Inclusion in social protection programs was more likely for older individuals with disabilities, those who were unmarried, widowed, divorced, or separated. Conversely, individuals with disabilities who had higher levels of education, resided in more affluent households, and lived in the Dhaka division were less likely to be included in these programs. Among children aged 0-17 years, the likelihood of inclusion in social protection programs was higher if they had more than one form of disability. These findings are robust, derived from an advanced statistical model applied to a large, nationally representative sample, and account for various confounding factors at the individual, household, and community levels. Notably, our findings highlight the inadequacy of social protection program inclusion for individuals with disabilities, with nearly two-thirds excluded from any such program.

This study highlights that social protection programs in Bangladesh include only 38% of individuals with disabilities, a figure that increases to 48% when considering their lifetime estimate. Within this group, 69% receive support through disability allowances tailored for them, while the remaining receive assistance from other sources, such as old age or widowed allowances. These findings underscore that a majority of persons with disabilities in Bangladesh remain excluded from social protection programs. Even among those included, roughly one-third are not integrated into programs designed to address their specific needs. This situation contrasts with other LMICs, like India and Pakistan, where the coverage of social protection programs for persons with disabilities is much higher (21%-63%) [33, 34]. The lower coverage of social protection program in Bangladesh is attributed to the government’s lower allocation of resources for vulnerable populations compared to the demand, despite an overall increase in the budget over the years [23]. For example, in the most recent 2022-2023 national budget in Bangladesh, only BDT 2978.91 core (2.8 billion USD) is allocated for disabled persons, with a monthly allowance of only 850 BDT (7.73 USD) [23]. Along with this lower funding availability, in many cases, this amount does not reach the intended recipients, with consistent evidence of incorrect beneficiary selection based on political identity, personal relationships with local leaders, and corruption among local leaders [7, 35]. This issue is compounded by poor governance, administrative complexity, and a lack of effective monitoring, further emphasizing the challenging conditions faced by people with disabilities in Bangladesh [27]. This indicates the challenging circumstances faced by people with disabilities in Bangladesh, with rising costs that many individuals with disabilities cannot afford, including increasing demand for healthcare services and regular visits to healthcare centers, which are often located in cities [8, 13]. Consequently, a significant proportion of individuals with disabilities (22-25%) struggle to meet their basic needs, including food and clothing [1].

Complexity also arises in national-level policies and programs that are designed to provide social support [7]. For instance, according to current policies and programs, a disabled child is only eligible for a disability allowance if they meet specific criteria: (1) they are aged six years or older, (2) they reside in a family with an annual income of less than 36,000 BDT (360 USD), and (3) they are diagnosed with [36, 37]. However, with rapid economic development, finding someone in Bangladesh with an annual income less than 36000 BDT has become increasingly unlikely. Furthermore, restricting eligibility to only a specific type of disabled child aged six years or older automatically excludes a significant number of children with disabilities [17]. A similar challenge exists for educational stipends, as children with disabilities are ineligible unless they are enrolled in government-recognized educational institutions [38]. This is often not the case, as they may attend schools not recognized by the government, like many disabled children who enroll in Madrasah education, which is often not government registered [5, 38].

We identified a decreased likelihood of inclusion in social protection programs among female individuals with disabilities and those with higher levels of education and wealth quintiles, as reported in other studies in LMICs [12, 30, 39]. Several factors could potentially contribute to these findings. For females with disabilities, cultural and societal factors may come into play. Gender biases and disparities in opportunities and decision-making could affect their access to social protection programs [40]. They also often face multiple layers of marginalization, due to both their gender and disability status, making them more vulnerable and potentially less likely to access social protection programs [2, 40]. Moreover, women with disabilities may contend with additional caregiving responsibilities, which can limit their ability to engage with these programs. Regarding individuals with higher levels of education and wealth quintiles, there may be a perception that they are more self-sufficient and less in need of social protection support [19, 41]. This assumption might lead to their exclusion from these programs, even though they may still require assistance [14, 39]. Additionally, individuals with higher levels of education may have a better understanding of their rights and eligibility for these programs, potentially leading to lower participation rates as they may seek alternative means of support [41, 42]. Similarly, those in higher wealth quintiles may have greater financial resources to address their needs independently, reducing their reliance on social protection programs [6, 15]. Furthermore, these disparities in inclusion may also result from variations in awareness and outreach efforts, with marginalized groups potentially receiving less information about available support [16]. Lastly, administrative complexities and issues with program accessibility may also contribute to these disparities, as individuals with greater access to information and resources may navigate these challenges more effectively [3].

Conversely, this study found that increasing age among disabled individuals, those engaged in white-collar occupations, students, and those unable to work, as well as unmarried, widowed, or divorced/separated individuals with disabilities, reported higher probabilities of inclusion in social protection programs [8]. These findings are consistent with research from other LMICs [4, 10, 30]. Several factors may underlie these findings [31]. As disabled individuals age increased, they may face greater challenges in terms of their physical or economic well-being, making them more vulnerable and increasing the likelihood of their inclusion in social protection programs [20]. Moreover, with the increasing age, they may be able to negotiate with the local leaders or program implementers to include them in the programs, otherwise they are mostly unidentified [7, 43]. Disabled persons working in white-collar occupations may have a stronger voice in advocating for their rights and accessing social protection programs, as well as a better understanding of their eligibility and how to navigate the system [15]. Students with disabilities may receive support through educational institutions or disability-specific scholarships, leading to a higher likelihood of inclusion in social protection programs [42]. Their status as students might make them more visible and more likely to receive assistance. Disabled individuals who are unable to work may face particularly acute financial hardships, prompting social protection programs to be more responsive to their needs and more inclined to include them [22, 39]. Similarly, unmarried, widowed, or divorced/separated individuals with disabilities may be seen as more vulnerable or in need of additional support, leading social protection programs to be more likely to encompass them. This increased inclusion could be driven by a combination of legal and policy considerations, which explicitly target these individuals as eligible beneficiaries, and a lack of alternative support systems for this demographic, making social protection programs a critical source of assistance [44]. Additionally, gender disparities and the societal recognition of their vulnerabilities may further contribute to their prioritization in such programs [45]. These insights highlight the complex interplay of individual characteristics, socioeconomic factors, and program design, emphasizing the need for tailored policies that address the unique needs and circumstances of persons with disabilities [26, 46]. These findings are also consistent with research from other LMICs and suggest that certain demographic and socioeconomic factors play a role in determining who is included in social protection programs [4, 26].

This study has several strengths and a few limitations. To the best of our knowledge, this is the first study in Bangladesh that explored the support received by persons with disabilities under the social support program at the national level and its correlates. It is based on a quite large sample collected through a nationally representative household survey, and recognized procedures were applied to measure social protection. Data were analyzed using comprehensive statistical modeling with a hierarchical structure of the data, and sampling weights were considered in all analyses. Therefore, the findings are robust and can be used in developing national-level policies and programs. However, the primary limitations of this study include the analysis of cross-sectional data, which limited our capacity to establish causality, and the findings were purely correlational. Data were collected through questions posed to the respondents with no opportunity for validation, indicating the possibility of recall bias, although any such bias is likely to be random. Furthermore, aside from the factors adjusted in the model, health and environmental variables can contribute to the onset of disability, making them important to be considered in the model. However, these data were not available in the survey, limiting our ability to do so. Nonetheless, despite these limitations, the findings of this study will contribute to national-level policy and program development.

## Conclusions

Only 38% of persons with disabilities reported being included in any social protection program within six months of the survey. This figure increased to 48% when considering support received at any time beyond six months. Disability allowances were the most commonly accessed form of social protection program, followed by old age allowances and assistance within the VGD/VGF programs. Inclusion in social protection programs was more likely for older individuals with disabilities and those who were unmarried, widowed, divorced, or separated. Conversely, individuals with disabilities who had higher levels of education and resided in wealthier households were less likely to be included in these programs. These findings highlight the inadequate coverage of social protection programs for persons with disabilities. Existing programs to support individuals with disabilities need to be strengthened to provide broader assistance to this vulnerable group.

## Declarations

## Data Availability

The datasets used and analysed in this study are available at the Bangladesh Bureau of Statistics.

## Acknowledgement

The authors thank the Bangladesh Bureau of Statistics (BBS)for granting access to the 2021 National Survey on Persons with Disabilities data.

## Authors’ contributions

Rahman MM and Rana MS designed the study, performed the data analysis, and wrote the first draft of this manuscript. Khan MN and Rahman MM critically reviewed and edited the previous versions of this manuscript. All authors approved this final version of the manuscript.

## Conflict of interests

The authors declare that they have no known competing financial interests or personal relationships that could have appeared to influence the work reported in this paper.

## Ethics approval and consent to participate

The survey protocol was reviewed and approved by the National Research Ethics Committee in Bangladesh. No additional ethical approval was required to conduct this study.

## Consent for publication

Not applicable

## Availability of data and materials

The datasets used and analysed in this study are available at the Bangladesh Bureau of Statistics.

## Conflict of interest

None.

## Funding

This research did not receive any specific grant from funding agencies in the public, commercial, or not-for-profit sectors.

## List of Abbreviations

PWD: Persons with disability
SSNP: Social Safety Net Program
WHO: World Health Organization
SBI: Short Birth Interval
LMICs: Low-and middle-income countries
BBS: Bangladesh Bureau of Statistics
mOR: Multinomial Odds Ratios; aOR, Adjusted Odds Ratios
AIC: Akaike Information Criteria
BIC: Bayesian Information Criteria.

## Notes

### Competing Interest Statement

The authors have declared no competing interest.

### Author Declarations

The survey protocol was reviewed and approved by the National Research Ethics Committee in Bangladesh.

